# SARS-CoV-2 serological findings and exposure risk among employees in school and retail after first and second wave COVID-19 pandemic in Oslo, Norway: a cohort study

**DOI:** 10.1101/2021.04.27.21256214

**Authors:** Anne-Mari Gjestvang Moe, Mina Eriksen, Tiril Schjølberg, Fred Haugen

## Abstract

**Background:** During initial phases of the coronavirus disease 2019 (COVID-19) pandemic, many workplaces were affected by closures and various preventive measures intended to limit infections. Here, we characterize and compare in an ambidirectional cohort study SARS-CoV-2 serology among Norwegian school employees and retail employees at baseline following the first epidemiological wave, and at follow-up after a second wave.

**Methods:** We enrolled a cohort of 238 school and retail employees after the first COVID-19 pandemic wave. Self-reported exposure history and serum samples were collected at 10 schools and 15 retail stores in Oslo, Norway, sampled at two time-points, baseline (May 18. to July 2. 2020) and follow-up (Jan 7. to Mar 17. 2021). SARS-CoV-2 antibodies targeting both spike and nucleocapsid were characterized by multiplex microsphere-based serological methods.

**Results:** At baseline, 6 enrolled workers presented with positive SARS-CoV-2 serology (3%; CI [1, 6]; P=0.019), which was significantly higher than the expected 1% prevalence in the general Oslo-population at this time-point. Five of the positive cases were retail employees. However, school and retail groups distributions at baseline were not significantly different as the number of seropositive observations were limited. Due to a school closure effectuated during the first wave, half of the school employees reported ≤2 days of physical workplace presence per week, while 65% of the retail employees reported ≥5 days per week. Eight months later, after passing a second epidemiological wave, school and retail groups presented 11 new seropositive cases altogether, but there was still no significant differences between the groups. Physical attendance at the workplace was similar between the groups during the second wave, but some preventive measures against viral transmission at workplaces were different. Self-reported virus diagnostics (RNA) for the same period were compared to the serological data obtained in this study, showing that all but one positive SARS-CoV-2 serological findings arising between baseline and follow-up had been diagnosed with virus testing.

**Conclusions:** After the first wave, distribution of SARS-CoV-2 positive serology was slightly higher than expected in a cohort of school and retail employees. Distribution of infection was not significantly different between the groups at baseline nor at follow-up, even though physical workplace attendance had been different. Nearly all new seropositive cases discovered in this study between baseline and follow-up, had already been diagnosed due to widespread virus testing during the second wave. This highlights the importance of extensive viral testing among workers.

## INTRODUCTION

The ongoing pandemic of COVID-19 (Coronavirus Disease -19), caused by the virus termed Severe Acute Respiratory Syndrome Coronavirus -2 (SARS-CoV-2), has had an unprecedented global impact on health and economy since it was declared a pandemic by the World Health Organization (WHO) March 11. 2020. At that point, the number of confirmed cases was approximately 120 thousand diagnosed cases worldwide; by the end of 2020 this number had risen to approximately 80 million confirmed cases (WHO 2020).

It has become clear that COVID-19 poses a threat to workers’ health. Some of the earliest reports of COVID-19 were from an occupational setting. Although somewhat disputed (Mallapaty 2021), the origins of the pandemic and some of the earliest cases reported were registered among fish market workers in Wuhan, China (Guo et al. 2020; Li et al. 2020). After initial global spread of the virus, health care workers were particularly affected. In April 2020, Italian health care workers amounted to 10% (approx. 12,000) of the nationwide total of registered cases (Chirico et al. 2020). In health care workers the adverse impact of COVID-19 has been documented (Carlsten et al. 2021), but vulnerability is not as well documented among other essential workers. Two aspects of COVID-19 among workers determine its impact: 1) individual risk factors like age, sex, preexisting conditions; and 2) workplace factors that influence SARS-CoV-2 exposure and eventually transmission at work (Leso et al. 2021). Since individual factors often are unmodifiable, preventive measures should be aimed towards viral exposures at the workplace (Carlsten et al. 2021): 1) exposure elimination (e.g. remote work, workplace closures, symptom checks); 2) engineering controls (e.g. shields, irradiation, hand hygiene); 3) personal protection equipment (PPE; e.g. masks, gowns, gloves); and 4) administrative controls (social distancing, staggered work schedules, behavioral requirements/training, surface cleaning/disinfection).

Diagnosis of SARS-CoV-2 in acutely or recently infected individuals usually involves molecular detection of viral RNA. Serological testing of SARS-CoV-2, on the other hand, has the advantage that it may be performed once viral RNA has been systemically cleared and symptoms have resolved (Parikh and Farnsworth 2021). Until now, studies relating to SARS-CoV-2 serology have largely focused on health care workers(Mhango et al. 2020). But there are some examples of studies of other groups relating to different sectors: meat processing industry workers (USA) (Herstein et al. 2021); factory workers (Germany and Croatia) (Jerković et al. 2021); university workers (Italy) (Grant et al. 2021); mine workers (Ivory Coast) (Milleliri et al. 2021); and airport workers (Colombia) (Malagón-Rojas et al. 2021). Literature reviews suggest heterogenous seroprevalence estimates among workers’ populations, ranging from 0.5% to 10% (Grant et al. 2021).

One study of SARS-CoV-2 infection in a single grocery retail store (USA) has been published (Lan et al. 2021), besides that, few studies targeting retail employees exist. Whilst some data on SARS-CoV-2 transmission is available from educational settings (Macartney et al. 2020), the main focus has been on children’s role in virus transmission (Lordan et al. 2020). Studies involving serology indicate that the majority of infections among children have been asymptomatic (Cohen et al. 2020; Snape and Viner 2020), but that children seem to spread the virus in the same manner as adults (Jones et al. 2020). The transmission of SARS-CoV-2 from asymptomatic cases in general remains unknown. Possibly, this can be clarified using serological methods.

From an occupational health perspective, we aimed to gather knowledge about SARS-CoV-2 infections in vulnerable groups of workers in Oslo, Norway. In this study, we aimed to evaluate the distribution of SARS-CoV-2 serology among school and retail employees against workplace factors pertaining to exposure risk and preventive measures. Our approach was to establish a cohort of school employees and retail employees at baseline after the first wave of the COVID-19 pandemic, and to follow the distribution of SARS-CoV-2 serology in these two potentially exposed populations.

## METHOD

### Study setting

According to the European Centre for Disease Prevention and Control and Norwegian Institute of Public Health (NIPH), the first case of SARS-CoV-2 in Norway was notified in the final week of February 2020 (NIPH 2020b). The first epidemiological wave in Norway (spring to early summer 2020) was largely affected by lockdown restrictions in society, starting March 12. 2020. During the first wave, closure of schools at all levels of education was implemented, whereas retail stores were allowed to stay open. Schools reopened in mid May 2020 and remained open through the fall semester. While school employees benefited from these mitigation efforts, retail workers continued to experience potential SARS-CoV-2 exposure risk as most of retail service remained open and the turnover increased, especially for construction and renovation products. After the first wave, by the end of June 2020, NIPH reported that there had been approximately 8,800 confirmed cases (165 cases/100,000 inhabitants) of COVID-19 in Norway since the start of the pandemic. Oslo county had the highest number of cases; 413 cases/100,000 inhabitants. Only 6.2% of the population had been tested at this point, and the number of actual cases was likely higher (NIPH 2020c). Based on mathematic modelling, NIPH still estimated that less than 1% of the population had been infected by the end of June (NIPH 2020a). This estimate was later confirmed by studies of seroprevalence. Based on testing for antibodies for SARS-CoV-2 in 900 samples of residual sera from 9 different laboratories in April/May 2020, the estimated seroprevalence in the Norwegian population was 1%. The highest estimated seroprevalence against SARS-CoV-2 was found in Oslo (4.3%) (Tunheim G. 2020). Based on the available data at that stage, we hypothesized a prevalence of 1% in the general population, including the non-diagnosed. After the passing of the second epidemiological wave, by the end of January 2021, the accumulated number of confirmed COVID-19 cases in Norway had reached 60,000 (1,142 cases per 100,000) (NIPH 2021a). Among these, around 16,000 were from Oslo county (NIPH 2021b). NIPH estimated that approximately 2% of the Norwegian population had been infected with SARS-CoV-2 so far. This is partly supported by a serological study in the Oslo area with 9500 participants randomly selected from two ongoing research studies (Norflu and MoBa), which shows that 1.4% of the participants had developed antibodies against SARS-CoV-2 (NIPH 2020d).

### Eligibility criteria, ethical considerations, and follow-up

Eligible participants were workers either male or female, age > 18 years. Informed consents were obtained from all participants of the study. Study approval was granted April 27. 2020 by the Regional Committee for Medical and Health Research Ethics in South-Eastern Norway (Reference number 134064). Data handling protocol was reviewed by NSD - Norwegian Centre for Research Data and approved April 29. 2020 (reference number 560357). All methods were performed in accordance with the relevant guidelines and regulations. Participants were recruited at 10 schools (N=112) and 15 retail stores (N=126), generally located around the eastern parts of Oslo, Norway in May 2020 and June 2020, respectively. Recruitment was done via email and by handing out written information as a flyer. Baseline collection of data and blood occurred in the period May 18. to July 2. 2020, whereas the follow-up occurred Jan 7. to Mar 17. 2021. A research team of 2-4 people from STAMI showed up at the schools and businesses during work hours and collected questionnaire data and biospecimens by appointment. Follow-up was accomplished through e-mail invitation. Participants were not presented with their serological test results from the baseline session until the follow-up session was finished.

### Blood sampling and Luminex serological assay

Venous blood was drawn from the cubital vein of the antecubital fossa. A flush tube (vacutainer K3EDTA) was used and immediately discarded prior to collecting a total of 20 mL blood from each participant, using BD Vacutainer^®^ Plasma Preparation Tubes (BD PPT^™^, 9.0 mg K_2_EDTA, BD, Franklin Lakes, USA) and BD Vacutainer^®^ Serum Separation Tubes (BD SST^™^ II, 5ml, BD, Franklin Lakes, USA). Tubes were immediately inverted several times. PPT were then centrifuged for 10 minutes at 2,100 × g, SST were left to coagulate at room temperature for minimum 30 minutes, before being centrifuged for 5 minutes at 2,100 x g. Aliquots of plasma and serum were stored at -80°C until analysis. All blood samples were analyzed using Luminex technology. Antigen detection was obtained using xMAP^®^ SARS-CoV-2 Multi-Antigen IgG Assay (RUO) (30-00127 Luminex), supplemented with xMAP SARS-CoV-2 IgG Control Kit (30-00129 Luminex). Prior to the analysis, the samples were diluted 1:400. The assay was performed according to the manufacturer’s instructions, and the following data analysis was conducted using software provided by the manufacturer (xMAP SARS-CoV-2, CN-SW77-01 Luminex).

The MFI signal for each sample was compared to threshold values defined by the manufacturer of the assay to define a positive or negative test result. A sample will only be defined as SARS-CoV-2 IgG positive if the nucleocapsid (N) target antigen level is above threshold and at least one other target antigen level [S1 spike subunit (S1) and/or the Receptor-binding domain (RBD)] is above threshold. A background control ensures that positive samples are not the result of non-specific binding. Besides using the control kit supplied by the manufacturer, an additional verification of the kit was performed. Samples collectedprior to the pandemic in 2019 were used as negative controls, supplemented with positive controls confirmed either by RNA diagnostic test or serological analysis performed by a different method. For two of the positive controls, we had a negative control from the same person.

### Questionnaire

In conjunction with blood sampling, participants were asked to answer a questionnaire about personal symptoms, previous COVID-19 diagnosis and workplace measures aimed to prevent viral transmission at the workplace. The questionnaire was completed on site via their smartphone or alternatively a tablet.

### Statistics

The null hypothesis was expressed as H0: pB=p0=1% and the alternative as H1: pB>p0, where p0 represents the cumulated proportion of infected in the general population at baseline (assumed to be 1%), and pB the proportion of infected in a subgroup of workers. Estimates showed that n=90 was needed in each group to detect a 10% deviation from p0 with a power of 80% and Type-I error rate of 5%. Statistical tests were used as indicated with Graphpad 9 (Binomial Test, Fisher’s exact test or Mann-Whitney test).

## RESULTS

We aimed to define the distribution of SARS-CoV-2 infections under the first pandemic wave in a cohort of two working populations. A cohort of potential eligible workers, male and female, age >18 years (N=238) were recruited from 10 schools (N=112) and 15 retail stores (N=126) in Oslo, Norway by written consent (Fig. 1). At baseline, 236 eligible participants completed questionnaires (1 withdrew consent, and 1 was excluded due to age <18), serology was obtained from 209 (27 were missing due to failed blood sampling). At follow-up 166 participants completed questionnaires and serology was obtained from 160 (N=6 cases of failed blood sampling, whereas 70 participants were lost from baseline to follow-up and did not respond to contact made by e-mail), i.e. the follow up rate was 70 % (81% for school, 60% for retail). Withdrawal from the study was not a source of participants loss.

**Fig 1:**
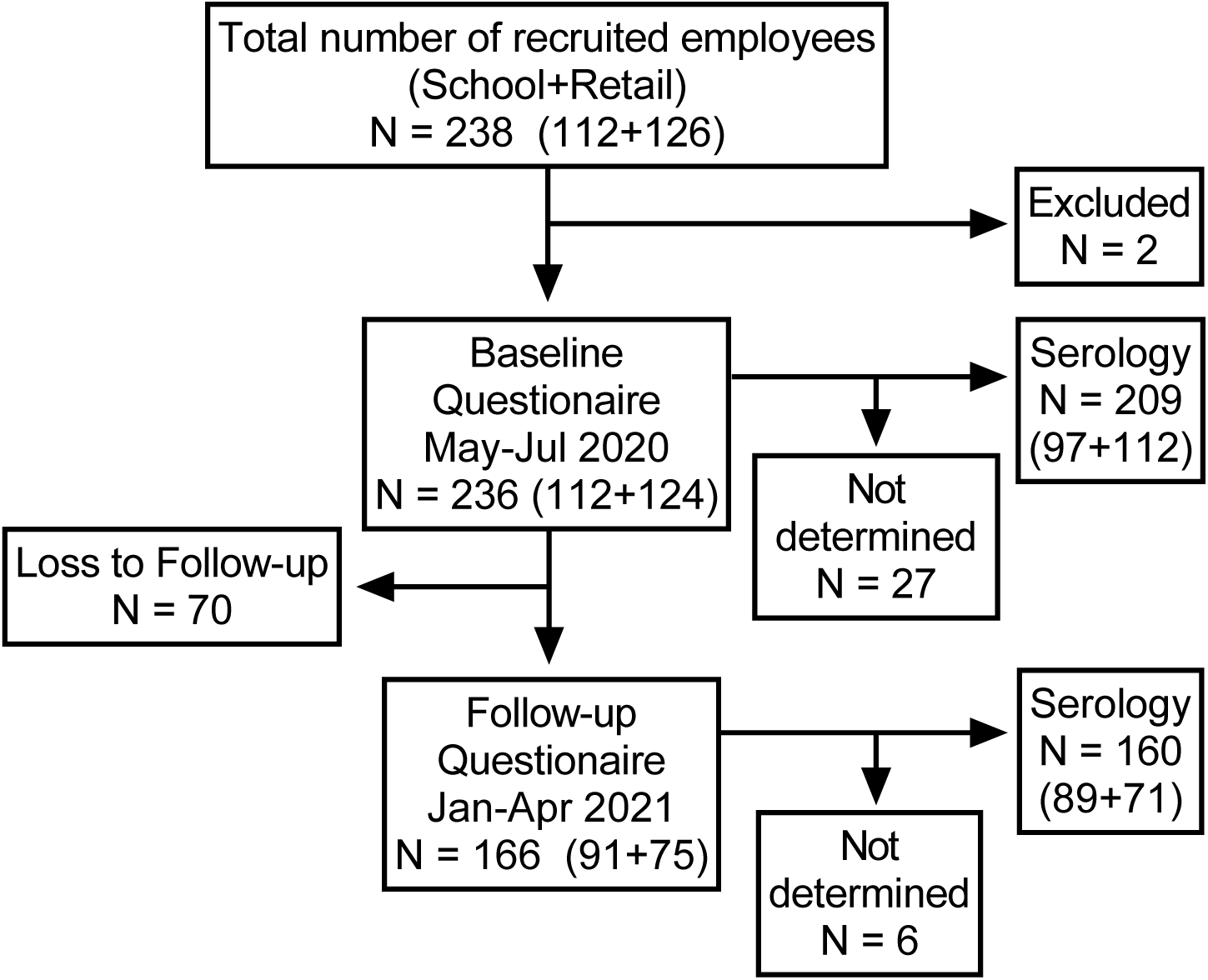
Schematic overview of participants in the study at baseline and follow-up.

Age was higher among employees in the school group than in the retail group. Sex distribution was skewed towards females in the school group, whereas more balanced in the retail group (Table 1). The school employees had pupils at different levels (ages); teaching levels were as follows: 22% level 1-4 (6-9 years); 16% level 5-7 (10-12 years); and 38% level 8-10 (13-15 years). In addition, 22% had no teaching, working in administration etcetera.

**Table 1.** Descriptive demographic baseline data of eligible employees in the cohort.

|  | School<br>(N=112) | Retail<br>(N=124) |
| --- | --- | --- |
| Mean age [years (min-max)] | 45 (21-68) | 36 (18-70) |
| Sex (M, F) | 27, 85 | 60, 64 |

First, we assessed the distribution of SARS-CoV-2 infections which had occurred during the first epidemiological wave in retrospect, using serological analyses of blood samples (N=209). For this period, we hypothesized that 1% of the general Oslo-population had been infected, which has been supported (NIPH 2020a). Multiplex microsphere-based SARS-CoV-2 serology of plasma samples showed that baseline distribution of seropositives within the whole workers’ cohort deviated from the anticipated level (3% incidence; CI [1, 6]; P=0.019) (Fig. 2A). Inspecting the two groups of workers separately, SARS-CoV-2 serology distribution among school employees after the first wave was not significantly different from the assumed distribution (1% incidence; CI [0.1, 6]; P=0.623), whereas the retail employees’ distribution was significantly different from the expected (4% incidence; CI [2, 10]; P=0.005). The distributions of positive test results between the two worker’s cohorts were compared directly with each other using Fisher’s Exact Test. The distribution of positive SARS-CoV-2 serology was not significantly different between the study groups (school employees relative to retail employees: OR 0.22; CI [0.02, 1.68]; P=0.219). Participants reported the average weekly number of days present at the workplace for the three months preceding the base line blood sample. As expected, school employees had significantly lower physical attendance at work than retail employees (Mann-Whitney test; P<0.001), due to school closure and remote work (Fig 2B). About 50% of school employees reported ≤2 days of physical workplace presence per week, whereas 65% of the retail employees reported ≥5 days.

**Fig 2:**
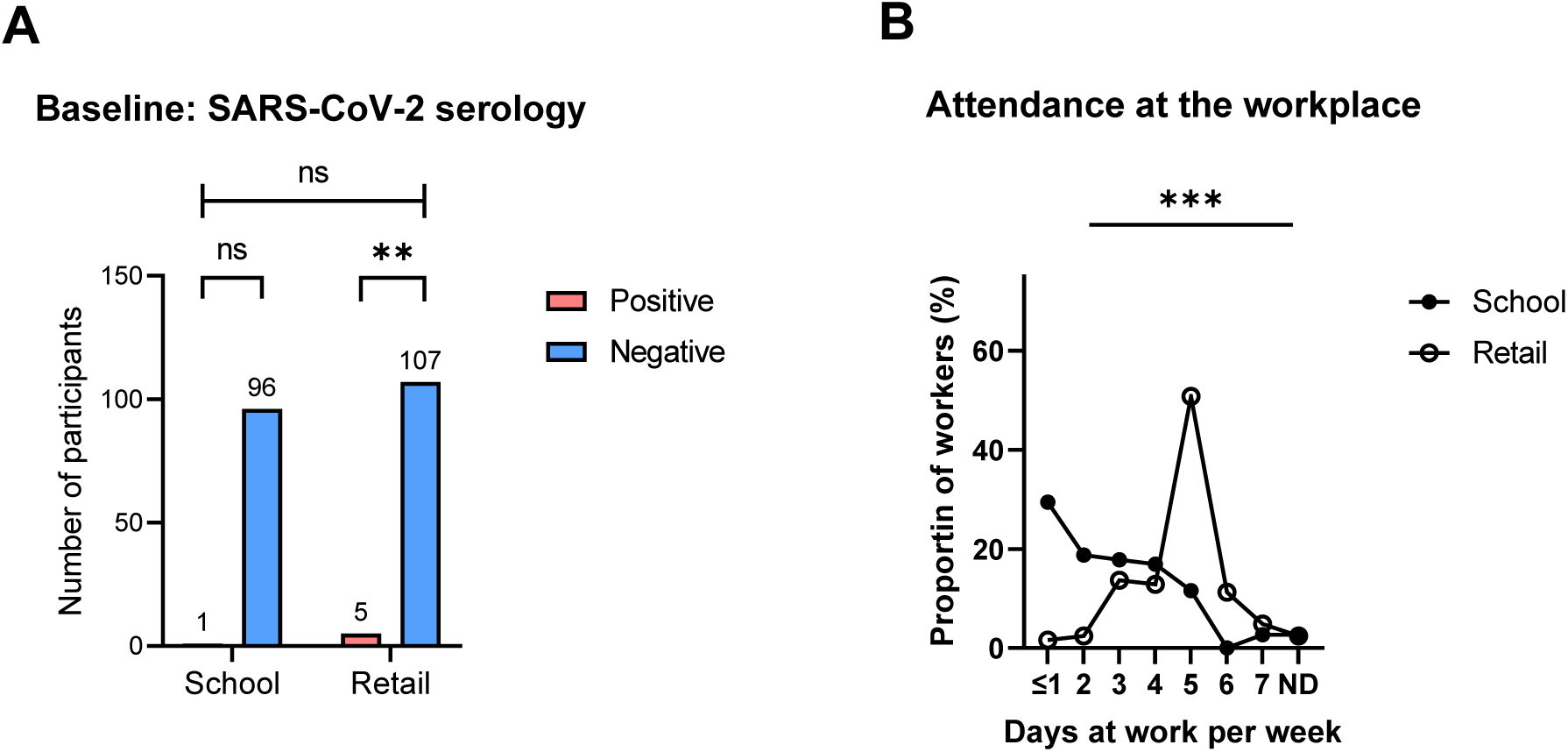
A) Baseline SARS-CoV-2 serology from a workers’ cohort sampled among school and retail employees in Oslo, Norway (in the period May 18.-July 2. 2020). The distribution of positive SARS-CoV-2 serology was compared between the study groups with Fisher’s exact test, not significant (ns). The unadjusted distribution of serology evaluated within each group was compared with the expected level of 1% seropositive in the general Oslo population using Binomial Test, **P≤0.01. B) Retrospective self-reported physical attendance at the workplace 3 months prior to the blood sampling at baseline; Mann-Whitney test between the school and retail groups, ***P≤0.001; ND, three non-responders in each group.

Second, by prospective design, 8 months later and after the passing of a second epidemiological wave, we identified individuals that had undergone COVID-19 infection by serological analyses of follow-up blood samples collected from the cohort (N=166; loss to follow-up=70). The number of SARS-CoV-2 seropositive cases at follow-up was increased from baseline (Fig 3A). School incidence was 8% over the 8-month follow-up period, whereas for retail incidence was 6%. Direct comparison of school and retail prevalence at follow-up, showed that there was no significant difference between the groups (OR 1.43; CI [0.4, 4.5]; P=0.756). At follow-up, participants (N=166) reported the average weekly number of days present at the workplace since the baseline sampling (Fig.3B). Ending of the universal school closure made school employees spend more time physically at the workplace. Now, 50% of school employees reported physical presence at their workplace ≥5 days per week. Employees in the retail group still spent more time at the workplace; 70% of the retail employees reported ≥5 days (Mann-Whitney test; P=0.0219) (Fig 3B).

**Fig 3:**
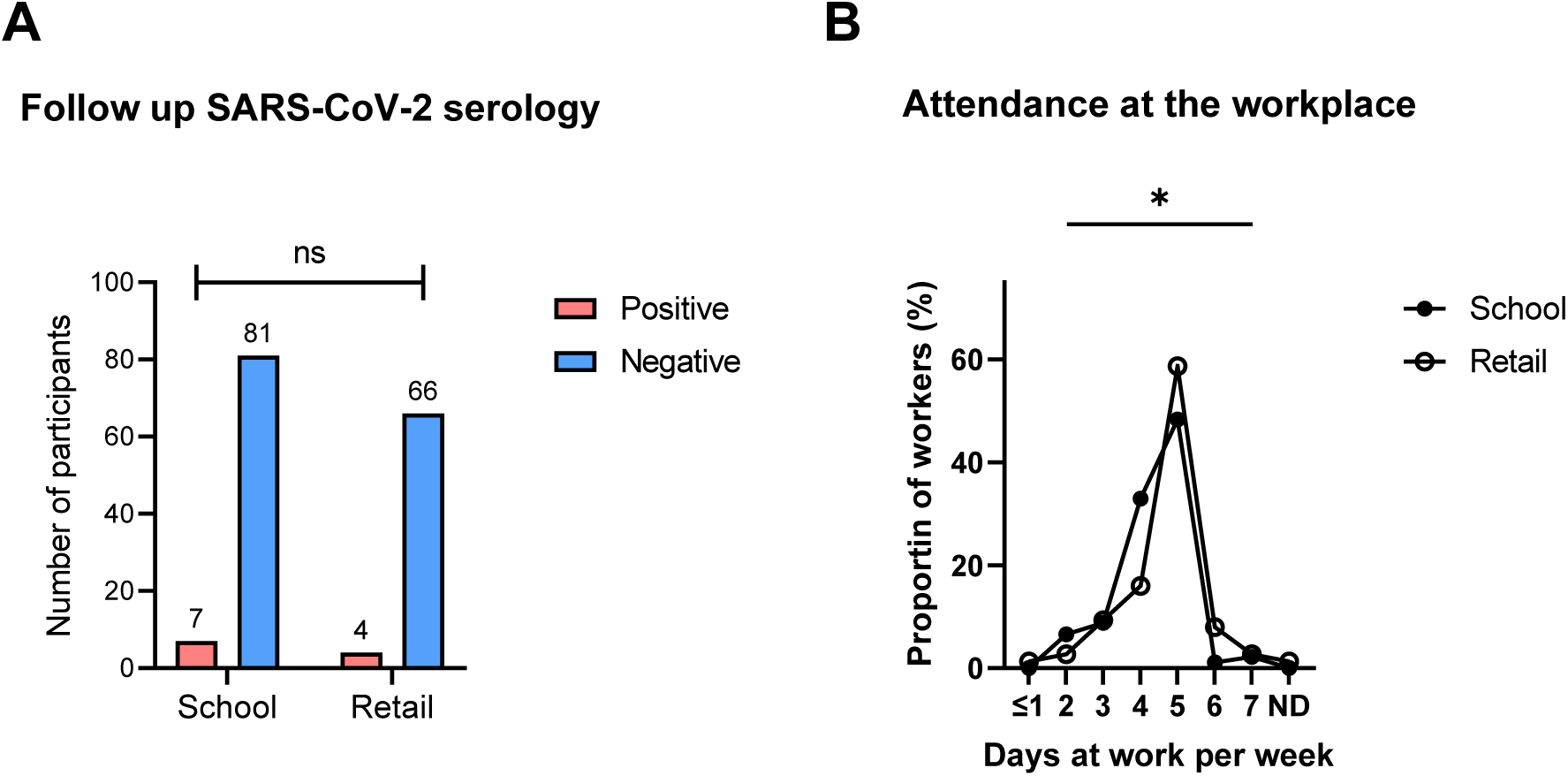
A) Follow-up SARS-CoV-2 serology in the cohort. The observed serological distributions in school and retail employees at 8 months follow-up were compared using two-sided Fisher’s exact test; ns, not significant. C) Retrospective self-reported physical attendance at the workplace 6 months prior to the blood sampling at follow-up. Mann-Whitney test between the school and retail groups, *P≤0.05; ND, one non-responder in the retail group.

Self-reported results of diagnostic SARS-CoV-2 RNA tests prior to baseline were aligned with the serological data to determine the level of undiagnosed SARS-CoV-2 infections in the cohort (Fig. 4). At baseline, 11 participants self-reported that they had been RNA-tested for ongoing infection; none of these tests had been positive. 196 participants self-reported that they had not been RNA-tested prior to baseline; among these, six individuals tested positive for SARS-CoV-2 serology. Virus testing became more available in the period between baseline and follow-up, hence at follow-up 114 participants self-reported that they had been RNA-tested for ongoing infection. Among these, 10 participants reported RNA-positive test, and all these were confirmed seropositive in our analyses at follow-up (Fig. 4). Three seropositive participants either received an RNA-negative result between baseline and follow-up (2 participants) or had not been RNA-tested (1 participant in the retail group). The data do not support the notion that undiagnosed SARS-CoV-2 infections were commonly occurring among workers in Oslo’s schools and retail stores during the second wave, although this was the rule during the first wave.

**Fig 4:**
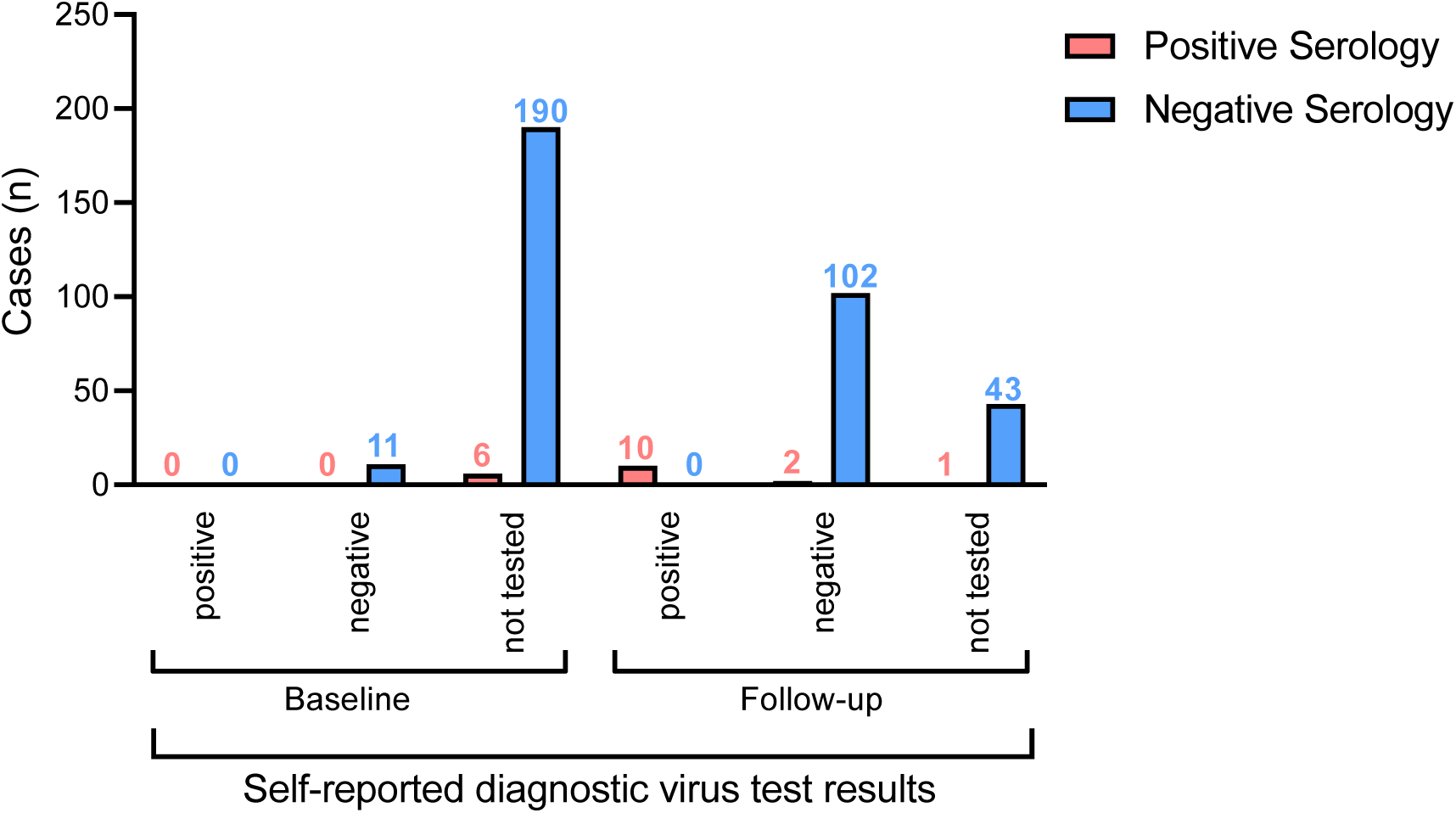
Self-reported virus test diagnostics (RNA) vs. serology (Ab) results among employees at baseline and follow-up. Participants were grouped according to self-reported RNA diagnostic results (lower panels) and its distribution was compared with the distribution of positive and negative serological results (bars). Numbers above bars indicate their value.

We aimed to map preventive measures taken against viral transmission at workplaces. Participating employees responded to a list of measures possibly implemented at their workplace (Table 2). Responses were different between school and retail employees: at baseline social distancing, contacts limitation, hand washing/disinfection, and surface washing/disinfection were more frequent among school-than retail employees. On the contrary, retail employees more often reported the use of personal protective equipment (PPE) and protective shields at baseline than school employees did. At follow-up, previous differences in social distancing and surface washing/disinfection were gone. A few participants reported symptom checks at work, and this was similar between the two groups. Similarly, a few also reported no measures; at baseline, this rate was higher in retail than among school employees.

**Table 2.**
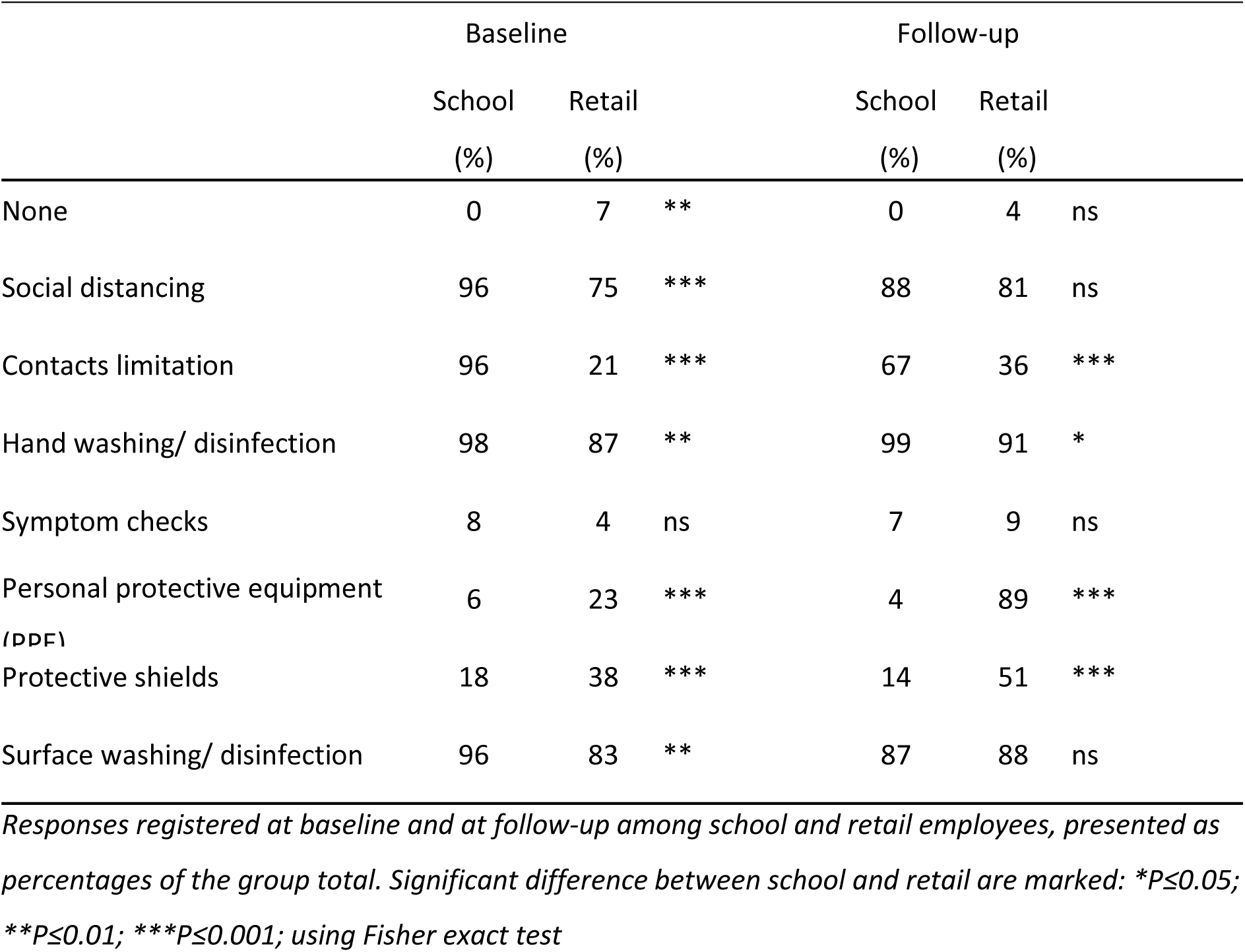
Preventive measures taken against viral transmission at workplaces.

## DISCUSSION

This study of SARS-CoV-2 serology in a cohort of Norwegian school and retail employees has given us new knowledge of the infectious distribution in two groups of employees with potentially high occupational exposure to the virus. Antibody measurements showed no significant difference in seropositive prevalence between the two groups at baseline after the first epidemic wave, nor at follow-up after the second wave. Compared to the assumed distribution of cases in the general population we noted at baseline that the number of cases in the cohort was higher than hypothesized. This was due to elevated occurrence of seropositive among retail employees in particular, whereas data on the school employee group showed more moderate levels. This difference in serology among the groups corresponded to a difference in attendance at the workplace during the first wave. A general school closure was enforced in Oslo during the first wave, whereas no general closures were imposed in retail. Matching serology with self-reported diagnostic test results showed low new incidence of undiscovered disease at follow-up. Serology uncovered a minor rate of undiagnosed cases in these populations of Oslo workers, hence the practice and level of diagnostic testing appeared to be adequate.

In this ambidirectional cohort study with baseline and follow-up serology, we aimed to observe SARS-CoV-2 infections, in retrospect from the origin of the pandemic until baseline (first wave), and prospectively from baseline until follow-up eight months later (second wave). The design has potential to uncover links between infection rates and exposure to occupation as a risk factor. New knowledge from this study confirms that effective alleviation kept COVID-19 transmissions relatively low in the Norwegian population during the first wave. However, low infection rates combined with the modest number of participants recruited in our study imposes limitations on the conclusions we can draw. The power estimates for the retrospective and prospective analyses to uncover the rate of undiagnosed cases in school and retail workers were based on scenarios made in April 2020 when this study was designed. Initially, we feared that the general population would face higher rates of disease transmission. In particular, we anticipated many infected workers in retail, as these individuals had exposure to potential transmission from customers and colleagues. Diagnostic testing capacity was insufficient at this stage, during the first wave, and many parameters were unknown. However, in contrast to the scenarios encountered in many other cities across the world, COVID-19 infection rates stayed relatively low in Oslo.

As the approach of this study was to visit workplaces to collect data on occupational health, authorization and organization at the workplaces was required. We achieved a feasible mode of data collection through recruiting participants in groups from a limited number of workplaces, 10 schools and 15 retail stores. These were an arbitrary selection of representative workplaces in Oslo, not a random sample of individuals drawn from the entire workers’ population, and thus might be affecting the study’s external validity. Sampling can affect results, as outbreaks of COVID-19 show clustering. On the other hand, with just a few workplaces to recruit from, more focus was put into encouraging participants, as well as streamlining sampling procedures to their work situation. This possibly leads to engaging participants that otherwise would not be motivated to participate, and is therefore likely to reduce selection bias. Potentially, loss to follow-up imposes limitations by affecting study validity, depending on how big of a loss, and how it is distributed. Here, we encountered 30 % loss to follow-up in the whole cohort, but most of the lost participants were lost from the group of retail employees. None of the participants withdrew from the study, but several were temporarily laid off, and some had started in other jobs, did not have the time, or did not answer our efforts to schedule follow-up sessions. Differential loss to follow-up can introduce bias that would lead to underestimation of prevalence, and therefore it is important to examine the possibility of an association between participant loss and contraction of COVID-19. Although we cannot exclude the possibility, we do not have observations to support a pattern of diminished work ability in the participants lost to follow-up. Based on the assumption that younger people experience more changing work markets, the lower mean age among retail employees could influence selectively on loss to follow-up. Otherwise, major differences introducing bias seem implausible between the participants lost to follow-up and the participants who completed the follow-up.

It is fundamental to this study that antibody measurements are reliable to detect individuals that have undergone SARS-CoV-2 infection. Studies show that most people infected with SARS-CoV-2 display an antibody response between day 10 and day 21 after infection, although detection in mild cases can take up to four weeks or more, and in some not detected at all. Post et al. concluded in their systematic review that SARS-CoV-2-specific IgM rises in the acute phase and peaks 2-5 weeks after disease onset, followed by a decline over 3-5 weeks until undetectable level in many cases. IgG peaks 3-7 weeks after disease onset, then plateaus or moderately declines at a persisting level (Post et al. 2020). Studies report a lasting level of antibody up to 6 to 12 months (Duysburgh et al. 2021), but the full duration and protective capacity of the immune response to SARS-CoV-2 is still unknown. Gudbjartsson et al. found a high seroprevalence (91,1%) in over 1000 persons recovered from a SARS-CoV-2 infection (Gudbjartsson et al. 2020). The same tendency has been confirmed in later publications (Dan et al. 2021). Serology, i.e. antibody tests, may therefore serve as a useful tool for detection of prior SARS-CoV-2 infections. In this study we measured SARS-CoV-2 antibodies targeting both spike and nucleocapsid using a multiplex microsphere-based serological method. Most assays on the market detect either antibodies directed against the nucleocapsid protein, the spike protein, or the receptor-binding domain (RBD) region of the spike protein (Parikh and Farnsworth 2021). With this multiplex assay we detect all three simultaneously, which is an advantage, given that high specificity is crucial, particularly with low prevalence.

A major finding of the present study was the scarcity of seropositive cases without COVID-19 diagnosis after the second wave. At follow-up only one seropositive participant was undiagnosed with COVID-19, the rest had been diagnosed earlier or detected at baseline in our study. The results from the second wave stood in contrast to the results from the first wave; at that point none of the seropositive reported that they had been diagnosed with COVID-19. Hence, with widespread testing nearly all cases are diagnosed. In the beginning of the COVID-19 pandemic in Norway, SARS-CoV-2 diagnostic RT-PCR test capacity was strictly limited. Patients admitted to hospital or other health care facilities with COVID-19 symptoms, health care professionals with symptoms, and symptomatic cases with underlying risk factors were prioritized for testing. Cases with known exposure and non-severe symptoms were not prioritized, and asymptomatic cases were not tested at all. Test capacity gradually improved during 2020. It is therefore likely that cases of COVID-19 infection, both asymptomatic and symptomatic, went undetected during the first wave, spring of 2020. A possible interpretation of our findings uncovers a low rate of asymptomatic cases. It is also possible that with enhanced awareness in the public and low threshold for testing, most cases are discovered. Oran and Topol approximated in their review of several cohorts tested for SARS-CoV-2 (up until May 2020), that the asymptomatic infection rate may be as high as 45% (Oran and Topol 2020). However, this review presented cross-sectional studies alongside longitudinal studies, and did not distinguish between asymptomatic and presymptomatic infection (subjects that are asymptomatic at the time of testing but will subsequently develop symptoms). In a recent review Buitrago-Garcia et al. address this issue and conclude that the overall estimate of the proportion of people who remain asymptomatic through the course of infection is 20% (Buitrago-Garcia et al. 2020). There have been reports that SARS-CoV-2-specific IgG levels in people with asymptomatic COVID-19 are significantly lower than in symptomatic patients, and that a larger portion of asymptomatic people become seronegative within 2-3 months, compared to symptomatic patients (Long et al. 2020). However, the results of this study have been questioned due to the fact that both symptomatic and asymptomatic patients received extensive anti-viral treatment, which may have affected the immune response (Koopmans and Haagmans 2020). In addition, Long et al. detected antibodies in 84% of the symptomatic patients, which may indicate that the antibody assay was not very sensitive (Long et al. 2020).

Another finding of our study was a higher distribution of seropositive than expected among retail employees, while this was not the case for school employees at baseline. Magnusson et al. have investigated the prevalence of COVID-19, based on diagnostic RT-PCR testing, in the Norwegian working population during the first and second wave of infection in Norway. During the first wave (February-July 2020), health care professionals and bus and taxi drivers had increased odds of COVID-19 infection compared to the average of the working population, while school workers had no, or even a reduced risk of COVID-19 when compared to the rest of the working population (Magnusson et al. 2021). This may be explained by reduced occupational exposure during the school closure in March, as well as a long summer vacation (June-July). This is reflected in our baseline data; the distribution of seropositive among school employees was not significantly different from the assumed distribution, and school employees reported a low level of physical attendance at work compared to retail employees. Our finding of a higher than expected distribution of seropositive among retail employees is not reflected in the study by Magnusson et al, but one must keep in mind that diagnostic testing was strictly limited during the first wave. The hypothesized 1% infected after the first wave was perhaps an underestimation of the true level, as studies of seroprevalence based on analyses of residual sera in the same period were estimated to be approximately 4% in Oslo (Tunheim G. 2020). Thus, it is also possible that the retail group in our study reflects the general distribution of SARS-CoV-2 infections in Oslo better, and that the incidence among school employees was low possibly due to the closure. During the second wave (July-November 2020) the odds of COVID-19 infection were higher for bartenders, waiters, transport conductors and travel stewards, according to Magnusson et al. Sales shop assistants and school workers were among the occupations with a moderately increased risk of COVID-19 during the second wave. However, the risk for school workers was up to double in Oslo county (Magnusson et al. 2021). In our study, we indeed find that at follow-up after the second wave, the distribution of seropositive in our cohort is higher than the assumed overall distribution. Stratification of new seropositive cases from baseline to follow-up indicated significantly increased new incidence among the school employees, while this was not the case for retail employees. Magnusson et al. argue that the supposed increased risk for school workers may be due to the fact that they are among the most frequently tested occupational groups (Magnusson et al. 2021). However, in our study we hardly find any undiagnosed cases in any of the groups studied during the second wave. Furthermore, it has not been clarified what school closure, and subsequent reopening, might contribute to COVID-19 control (Viner et al. 2020).

We conclude that seroprevalence in the study cohort was higher than the assumed level after the first wave, mainly caused by a high incidence among retail employees relative to school employees, and possibly linked to workplace attendance/remote work during the first wave. In contrast, attendance at the workplace during the period from baseline to follow-up (second wave) was similar between the two groups. Distribution of positive serology was not statistically different between school and retail employees at baseline nor at follow-up. Among the new cases between baseline and follow-up, we could hardly find any that were undiagnosed; hence the level of diagnostic testing seemed adequate.

## Data Availability

All of the data is included in the paper

## REFERENCES

Buitrago-Garcia D, Egli-Gany D, Counotte MJ, Hossmann S,Imeri H, Ipekci AM, Salanti G, and Low N (2020) Occurrence and transmission potential of asymptomatic and presymptomatic SARS-CoV-2 infections: A living systematic review and meta-analysis. PLoS Med. 17: e1003346. doi: 10.1371/journal.pmed.1003346

Carlsten C, Gulati M, Hines S, Rose C, Scott K, Tarlo SM, Torén K, Sood A, et al. (2021) COVID-19 as an occupational disease. Am J Ind Med. 64: 227–237. doi: 10.1002/ajim.23222

Chirico F,Nucera G, and Magnavita N (2020) COVID-19: Protecting Healthcare Workers is a priority. Infect Control Hosp Epidemiol. 1–4. doi: 10.1017/ice.2020.148

Cohen R, Jung C, Ouldali N, Sellam A, Batard C, Cahn-Sellem F, Elbez A, Wollner A, et al. (2020) Assessment of spread of SARS-CoV-2 by RT-PCR and concomitant serology in children in a region heavily affected by COVID-19 pandemic. medRxiv. 2020.2006.2012.20129221. doi: 10.1101/2020.06.12.20129221

Control ECfDPa. https://www.ecdc.europa.eu/en/covid-19/latest-evidence/immune-responses. doi:

Dan JM, Mateus J, Kato Y, Hastie KM, Yu ED, Faliti CE, Grifoni A, Ramirez SI, et al. (2021) Immunological memory to SARS-CoV-2 assessed for up to 8 months after infection. Science. 371: eabf4063. doi: 10.1126/science.abf4063

Duysburgh E, Mortgat L, Barbezange C, Dierick K, Fischer N, Heyndrickx L, Hutse V, Thomas I, et al. (2021) Persistence of IgG response to SARS-CoV-2. Lancet Infect Dis. 21: 163–164. doi: 10.1016/S1473-3099(20)30943-9

Grant R, Dub T, Andrianou X, Nohynek H, Wilder-Smith A, Pezzotti P, and Fontanet A (2021) SARS-CoV-2 population-based seroprevalence studies in Europe: a scoping review. BMJ Open. 11: e045425. doi: 10.1136/bmjopen-2020-045425

Gudbjartsson DF, Norddahl GL, Melsted P, Gunnarsdottir K, Holm H, Eythorsson E, Arnthorsson AO,Helgason D, et al. (2020) Humoral Immune Response to SARS-CoV-2 in Iceland. N Engl J Med. 383: 1724–1734. doi: 10.1056/NEJMoa2026116

Guo L, Ren L, Yang S, Xiao M, Chang D, Yang F, Dela Cruz CS, Wang Y, et al. (2020) Profiling Early Humoral Response to Diagnose Novel Coronavirus Disease (COVID-19). Clin Infect Dis. doi: 10.1093/cid/ciaa310

Herstein JJ, Degarege A, Stover D, Austin C, Schwedhelm MM, Lawler JV, Lowe JJ, Ramos AK, et al. (2021) Characteristics of SARS-CoV-2 Transmission among Meat Processing Workers in Nebraska, USA, and Effectiveness of Risk Mitigation Measures. Emerg Infect Dis. 27: 1032–1038. doi: 10.3201/eid2704.204800

Jerković I, Ljubić T, Bašić Ž, KruŽić I, Kunac N, Bezić J, Vuko A, Markotić A, et al. (2021) SARS-CoV-2 Antibody Seroprevalence in Industry Workers in Split-Dalmatia and šibenik-Knin County, Croatia. J Occup Environ Med. 63: 32–37. doi: 10.1097/jom.0000000000002020

Jones TC, Mühlemann B, Veith T, Biele G, Zuchowski M, Hofmann J, Stein A, Edelmann A, et al. (2020) An analysis of SARS-CoV-2 viral load by patient age. medRxiv. 2020.2006.2008.20125484. doi: 10.1101/2020.06.08.20125484

Koopmans M, and Haagmans B (2020) Assessing the extent of SARS-CoV-2 circulation through serological studies. Nature Medicine. 26: 1171–1172. doi: 10.1038/s41591-020-1018-x

Lan F-Y,Suharlim C,Kales SN, and Yang J (2021) Association between SARS-CoV-2 infection, exposure risk and mental health among a cohort of essential retail workers in the USA. Occupational and Environmental Medicine. 78: 237–243. doi: 10.1136/oemed-2020-106774

Leso V, Fontana L, and Iavicoli I (2021) Susceptibility to Coronavirus (COVID-19) in Occupational Settings: The Complex Interplay between Individual and Workplace Factors. International Journal of Environmental Research and Public Health. 18: 1030. doi: doi: https://www.mdpi.com/1660-4601/18/3/1030

Li Q, Guan X, Wu P, Wang X, Zhou L, Tong Y, Ren R, Leung KSM, et al. (2020) Early Transmission Dynamics in Wuhan, China, of Novel Coronavirus-Infected Pneumonia. N Engl J Med. 382: 1199–1207. doi: 10.1056/NEJMoa2001316

Long Q-X, Tang X-J, Shi Q-L, Li Q,Deng H-J, Yuan J, Hu J-L, Xu W, et al. (2020) Clinical and immunological assessment of asymptomatic SARS-CoV-2 infections. Nature Medicine. 26: 1200–1204. doi: 10.1038/s41591-020-0965-6

Lordan R,FitzGerald GA, and Grosser T (2020) Reopening schools during COVID-19. Science. 369: 1146. doi: 10.1126/science.abe5765

Macartney K, Quinn HE, Pillsbury AJ, Koirala A, Deng L,Winkler N, Katelaris AL, O’Sullivan Mvn, et al. (2020) Transmission of SARS-CoV-2 in Australian educational settings: a prospective cohort study. Lancet Child Adolesc Health. 4: 807–816. doi: 10.1016/S2352-4642(20)30251-0

Magnusson K, Nygård K, Methi F,Vold L, and Telle K (2021) Occupational risk of COVID-19 in the 1<sup>st</sup> vs 2<sup>nd</sup> wave of infection. medRxiv. 2020.2010.2029.20220426. doi: 10.1101/2020.10.29.20220426

Malagón-Rojas JN, Rubio V, and Parra-Barrera E (2021) Seroprevalence and seroconversions for SARS-CoV-2 infections in workers at Bogota Airport, Colombia 2020. J Travel Med. doi: 10.1093/jtm/taab006

Mallapaty S (2021) What’s next in the search for COVID’s origins. Nature. doi: 10.1038/d41586-021-00877-4

Mhango M, Dzobo M, Chitungo I, and Dzinamarira T (2020) COVID-19 Risk Factors Among Health Workers: A Rapid Review. Saf Health Work. 11: 262–265. doi: 10.1016/j.shaw.2020.06.001

Milleliri JM, Coulibaly D, Nyobe B, Rey JL, Lamontagne F, Hocqueloux L, Giaché S, Valery A, et al. (2021) SARS-CoV-2 Infection in Ivory Coast: A Serosurveillance Survey among Gold Mine Workers. Am J Trop Med Hyg. doi: 10.4269/ajtmh.21-0081

NIPH. 2020a. ‘COVID-19-epidemien: Kunnskap, situasjon, prognose, risiko og respons i Norge etter uke 26’. https://www.fhi.no/contentassets/c9e459cd7cc24991810a0d28d7803bd0/vedlegg/covid-19-epidemien---kunnskap-situasjon-prognose-risiko-og-respons-i-norge-etter-uke-26-01.07.2020.pdf.

NIPH. 2020b. ‘COVID-19 ukesrapport-uke 11’. https://www.fhi.no/contentassets/8a971e7b0a3c4a06bdbf381ab52e6157/vedlegg/forste-halvar--2020/2020-03-19-ukerapport-covid-19.pdf.

NIPH. 2020c. ‘COVID-19 Ukesrapport -uke 26’. https://www.fhi.no/contentassets/8a971e7b0a3c4a06bdbf381ab52e6157/vedlegg/andre-halvar--2020/2020.07.01-ukerapport-uke-26-covid-19-pdf.pdf.

NIPH. 2020d. ‘Hvor mange har vært smittet med koronavirus i Oslo og omegn?’. https://www.fhi.no/studier/prevalensundersokelser-korona/resultat---moba/

NIPH. 2021a. ‘COVID-19 Ukerapport – uke 3’. https://www.fhi.no/contentassets/8a971e7b0a3c4a06bdbf381ab52e6157/vedlegg/2021/ukerapport-for-uke-3-2021.pdf.

NIPH. 2021b. ‘Statistikk om koronavirus og covid-19’. https://www.fhi.no/sv/smittsomme-sykdommer/corona/dags--og-ukerapporter/dags--og-ukerapporter-om-koronavirus/#table-container-23157425

Oran DP, and Topol EJ (2020) Prevalence of Asymptomatic SARS-CoV-2 Infection : A Narrative Review. Ann Intern Med. 173: 362–367. doi: 10.7326/M20-3012

Parikh BA, and Farnsworth CW (2021) Laboratory evaluation of SARS-CoV-2 in the COVID-19 pandemic. Best Pract Res Clin Rheumatol. 35: 101660. doi: 10.1016/j.berh.2021.101660

Post N, Eddy D, Huntley C, van Schalkwyk MCI, Shrotri M, Leeman D, Rigby S, Williams SV, et al. (2020) Antibody response to SARS-CoV-2 infection in humans: A systematic review. PLoS One. 15: e0244126. doi: 10.1371/journal.pone.0244126

Snape MD, and Viner RM (2020) COVID-19 in children and young people. Science. 370: 286–288. doi: 10.1126/science.abd6165

Tunheim G. K AB., Rø, G., Steens, A., Hungnes O., Lund-Johansen, F., Tran, T., Andersen JT., Vaage, JT.. (2020) “Seroprevalence of SARS-CoV-2 in the Norwegian population measured in residual sera – analyses collected in April/May 2020 and August 2019.” In.: NIPH.

Viner RM, Russell SJ, Croker H, Packer J, Ward J, Stansfield C, Mytton O, Bonell C, et al. (2020) School closure and management practices during coronavirus outbreaks including COVID-19: a rapid systematic review. Lancet Child Adolesc Health. 4: 397–404. doi: 10.1016/S2352-4642(20)30095-X

WHO. 2020. ‘COVID-19 Weekly Epidemiological Update ‘. https://www.who.int/publications/m/item/weekly-epidemiological-update29---december-2020.

